# Estimating the effect of circulating vitamin D on body mass index: a Mendelian randomization study

**DOI:** 10.1101/2023.08.01.23293487

**Authors:** Minal Chadha, Joshua A. Bell, Eleanor Sanderson

**Affiliations:** Population Health Sciences, Bristol Medical School, University of Bristol, Bristol, UK; MRC Integrative Epidemiology Unit, University of Bristol, Bristol, UK

## Abstract

**Background:** Numerous observational studies have shown an association between higher circulating 25 hydroxyvitamin D (vitamin D) and lower body mass index (BMI). Whether this represents a causal effect remains unclear. Mendelian randomization (MR) is an approach to causal inference that uses genetic variants as instrumental variables to estimate the effect of exposures on outcomes of interest. MR estimates are not biased by confounding, reverse causation and other biases in the same way as conventional observational estimates. In this study, we used MR with new data on genetic variants associated with vitamin D to estimate the effect of vitamin D on BMI.

**Methods:** We selected single nucleotide polymorphisms (SNPs) which were associated with vitamin D in a recent large genome-wide association study (GWAS) at genome-wide significance as instruments for vitamin D. We used inverse variance weighted models and further assessed individual SNPs that showed evidence of an effect, and biologically informed SNPs located in genetic regions previously associated with vitamin D, for associations with other traits at genome-wide significance, using Wald ratio estimation.

**Result:** Our main results showed no evidence of an effect of vitamin D on BMI (estimated standard deviation change in BMI per standard deviation change in vitamin D: -0.003, 95% confidence interval [-0.06, 0.06]). This was also supported by pleiotropy robust sensitivity analyses. Individual SNPs that showed evidence of an effect of vitamin D on either lower or higher BMI were strongly associated with numerous other traits suggesting high levels of horizontal pleiotropy. Biologically informed SNPs showed no evidence of a causal effect of vitamin D on BMI and showed substantially less evidence of pleiotropic effects.

**Conclusion:** The observed association between vitamin D and BMI is unlikely to be due to a causal effect of vitamin D on BMI. We also show how additional evidence can be incorporated into an MR study to interrogate individual SNPs for potential pleiotropy and improve interpretation of results.

## Introduction

Nearly 2 billion adults are thought to be above a healthy weight body mass index (BMI) ≥25kg/m^2^), including more than 650 million adults with obesity (BMI >=30kg/m^2^)^1^. Rates of obesity have been increasing over recent years^2^ and obesity has been causally linked to many adverse health outcomes including type 2 diabetes, osteoarthritis, cardiovascular disease and increased mortality.^1,3,4^ However, BMI is hard to modify, with many interventions showing limited success, and obesity is difficult to reverse through lifestyle interventions once established.^5,6^ Numerous observational studies have shown an association between higher circulating 25 hydroxyvitamin D (vitamin D) and lower BMI and obesity-related diseases.^7-9^ The question of whether increasing vitamin D levels cause lower BMI is therefore one of substantial public interest, as it potentially presents a relatively easy way to intervene to lower BMI. However, conventional observational studies are limited in their ability to infer the causality or direction of this association and may be biased by unobserved confounding.^10^

Mendelian randomization (MR) is a form of instrumental variable (IV) estimation that uses genetic variants (single nucleotide polymorphisms (SNPs)) known to associate with an exposure of interest as instruments to estimate the causal effect of that exposure on an outcome.^11-13^ MR relies on the three key IV assumptions for the effect estimates obtained to be reliable: first, that the instruments must be strongly associated with the exposure; second, that there must be no unmeasured confounders of the instruments-outcome associations; and third, that there must be no effect of the instruments on the outcome that is not mediated via the exposure. These assumptions are illustrated in **Figure 1**. As genetic variants are inherited randomly at birth, they are less prone to confounding by lifestyle and socioeconomic factors and reverse causation^14^. According to classical twin studies, 50-60% of the variability in the concentration of vitamin D is explained by genetic factors indicating that it is a highly heritable trait.^15,16^ In this study, we performed a two-sample summary data MR study to estimate the causal effect of vitamin D levels on BMI.

**Figure 1.**
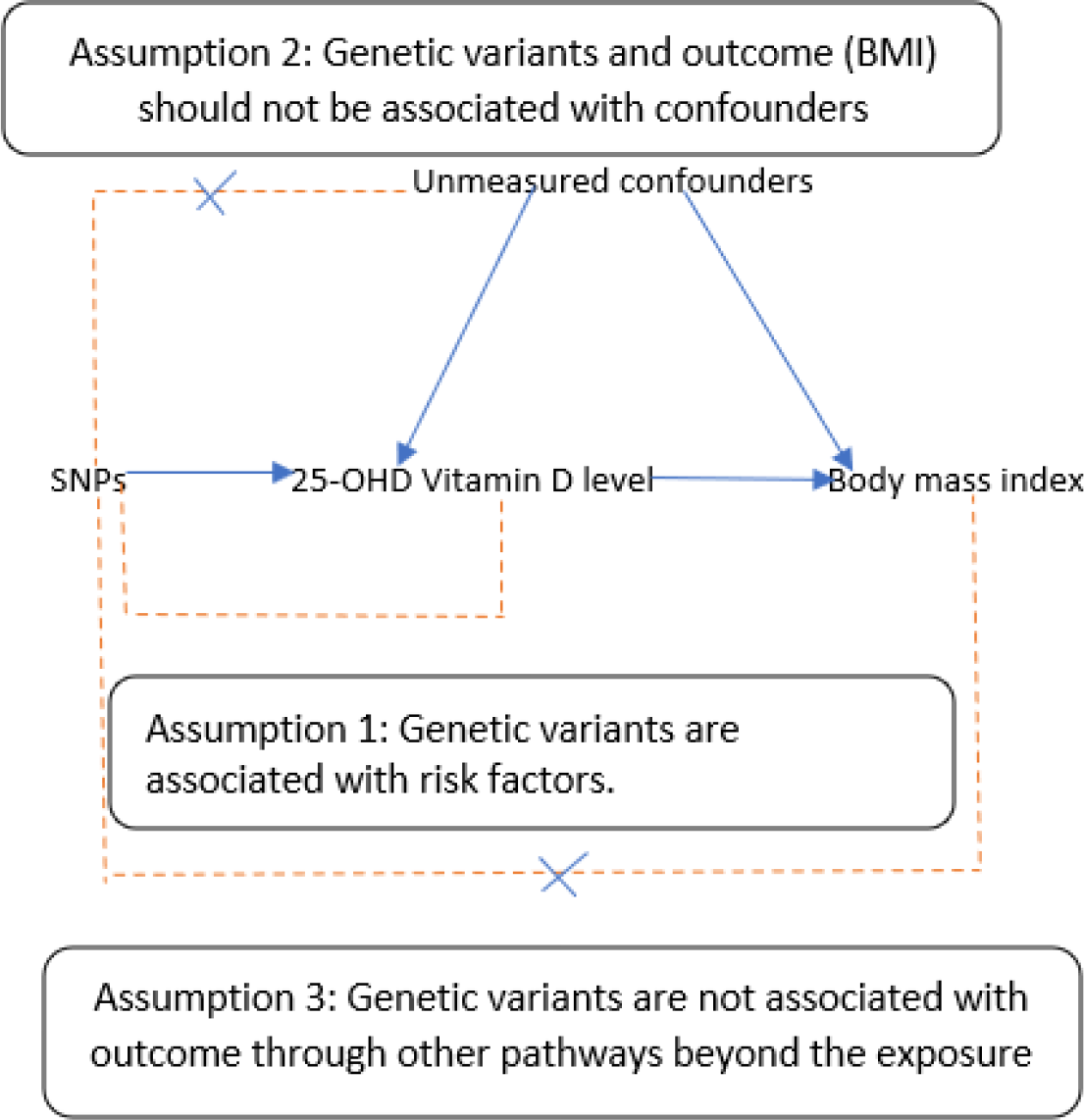
Study aim & assumptions. Study aims to estimate the effect of 25-OHD 25 hydroxyvitamin D level on BMI using genetic instruments of 25 hydroxyvitamin D. As explained in the figure, three assumptions of Mendelian randomization should be fulfilled to give unbiased results are explained.

This topic has been previously considered by an MR study which found no evidence of a causal effect of vitamin D on BMI.^17^ However, the previous study used only 4 SNPs as instruments for vitamin D, which were combined in an unweighted allele score due to weights for each SNP being unavailable, and had a sample size for analysis of up to 42,024 individuals. The debate in the literature about the potential causality and direction of the association between vitamin D and adiposity is ongoing.^18,19^ A new genome-wide association study (GWAS) of vitamin D has recently become available, based on a much larger sample than previous studies.^20,21^ This GWAS identified many more SNPs associated with vitamin D, 69 independent loci compared to the 4 previously available. These can then be used as instruments in a two-sample summary data MR study, along with a large recent GWAS of BMI for the outcome. Increasing the number of SNPs used in MR estimation increases the power to detect a causal effect if there is one but also raises the concern that some of these variants will be pleiotropic, violating the third IV assumption and biasing the overall effect estimate. Alongside applying standard pleiotropy-robust MR methods, we therefore explored the potential for pleiotropy in individual SNPs that showed evidence of a causal effect of vitamin D on BMI by examining their associations with other adiposity-related traits, and by examining biologically informed SNPs located in gene regions for vitamin D^22,23^.

## Methods

### Data sources

For our exposure data, we used summary statistics from a recent vitamin D GWAS of 443,734 individuals of European ancestry, consisting of 401,406 white British UK Biobank participants meta-analysed with a previous study of 42,274 Europeans.^20^ We selected SNPs that were associated with vitamin D at genome-wide significance (p < 5 ×10^−8^) and were pruned to exclude any which were in linkage disequilibrium with R^2^ > 0.001. For our outcome data, we extracted the SNP-outcome association for those SNPs identified as associated with the exposure from a recent large GWAS of BMI in individuals of white European ancestry.^24^ In this GWAS, BMI had been standardised and so the SNP-outcome associations extracted reflect a standard deviation (SD) change in BMI. This GWAS had a sample size of 681,275 and partially overlapped with the data used to generate the vitamin D GWAS used for our exposure; 456,426 of these individuals were from UK Biobank, representing about 67% of the sample. We harmonised the exposure and outcome summary data and excluded palindromic SNPs with a minor allele frequency >0.4.

### Study design

We conducted a two-sample MR study using inverse variance weighting (IVW) estimation.^25,26^ To test instrument strength, we calculated the mean F-statistic for the SNP-exposure association. We used a range of approaches to identify potential horizontal pleiotropy. Firstly, we obtained pleiotropy-robust effect estimates using MR Egger^27^, weighted mode^28^ and weighted median^29^ estimators. Each of these estimators are valid under more relaxed assumptions about the form of any pleiotropy. We then calculated the individual SNP-level effect estimates using the Wald ratio. We compared the proportion of these individual estimates that showed evidence of a causal effect to the number that would be expected under the null hypothesis of no causal effect. Evidence of a causal effect of vitamin D on BMI in the per-SNP analysis was defined as a p-value for the individual SNP Wald ratio of less than 0.05 divided by the total number of SNPs considered, to account for multiple testing. For each SNP that showed evidence of a causal effect, we conducted a phenome-wide association study (PheWAS) by looking up all other traits that SNP was associated with at genome-wide significance using the OpenGWAS platform.^30^ As a comparison, we identified SNPs in the GWAS that had been previously identified as being associated with vitamin D and were located in gene regions thought to be biologically relevant for circulating vitamin D. For these SNPs, we conducted the same SNP-lookup approach to find other traits associated with them.

Although MR is robust to reverse causation, if the true direction of effect is from the outcome to the exposure, and if any of the SNPs selected as instruments for the exposure are associated with the outcome, then the MR effect estimates will be biased. To explore whether this causes bias in our MR effect estimates, we applied Steiger filtering^31^ between our exposure and outcome data to remove any SNPs that explained more variation in BMI than vitamin D. Finally, we conducted an additional MR estimation in the reverse direction to estimate the effect of BMI on vitamin D. For this analysis, we used the same GWAS studies but selected all SNPs that were associated with BMI at genome-wide significance and applied the same summary-data MR methods for estimation as in our main analysis. The same harmonization and pruning steps were applied to these SNPs as before.

All analyses were carried out using R (version 4.2.2) and the TwoSampleMR package.^32^

## Results

69 SNPs were identified as being independently associated with vitamin D from the GWAS data. After harmonization with our outcome data, 68 SNPs were available to be used for the MR analysis. The mean F-statistic across those 68 SNPs was 142.9, which is greater than the approximated critical value of 10, suggesting that our MR estimates are unlikely to be biased by weak instruments.

Results from our MR analysis are given in **Table 1**. IVW estimation showed no evidence of an effect of vitamin D on BMI; the effect of an SD increase in vitamin D on an SD change in BMI was -0.003 SD, 95% confidence interval (CI): [-0.06, 0.06]. Results from the pleiotropy-robust methods were consistent with this result with only weighted mode showing any evidence of a small causal effect of vitamin D on BMI (beta: 0.026 SD, 95% CI: [0.01, 0.05]).

**Table 1:** MR estimates for the effect of vitamin D on BMI.

|  | Beta | SE | P-value | 95% C.I. |
| --- | --- | --- | --- | --- |
| *Without Steiger filtering* |  |  |  |  |
| IVW | -0.003 | 0.030 | 0.92 | [-0.06, 0.06] |
| MR-Egger | 0.047 | 0.047 | 0.33 | [-0.05, 0.15] |
| Weighted median | 0.011 | 0.014 | 0.44 | [-0.01, 0.03] |
| Weighted mode | 0.026 | 0.012 | 0.03 | [0.01, 0.05] |
| *With Steiger filtering applied* |  |  |  |  |
| IVW | -0.001 | 0.027 | 0.96 | [-0.03 0.02] |
| MR-Egger | 0.043 | 0.042 | 0.31 | [0.00 0.08] |
| Weighted median | 0.018 | 0.014 | 0.19 | [-0.01 0.05] |
| Weighted mode | 0.025 | 0.012 | 0.05 | [0.00 0.05] |

MR estimates for the effect of vitamin D on BMI. Number of SNPs: 68 – main analysis, 66 after Steiger filtering. $\beta$ : Estimated SD change in BMI per SD increase in vitamin D, SE: standard error, 95 % C.I.: 95% confidence interval.

As well as calculating the overall MR effect estimates we calculated the individual Wald ratios for each SNP. These are given in **Figure 2** and **Supplementary Table 1**. A number of these showed evidence for a causal effect of vitamin D on BMI. Using a p-value corrected for the number of tests done using Bonferroni correction (p=0.05/68 = 0.00074) as a cut-off for SNPs to explore further, we selected all SNPs that showed evidence of a causal effect for follow up in our PheWAS analysis. This resulted in 15 out of the 68 SNPs being selected, substantially more than would be expected by chance. Results from the PheWAS are summarised in **Table 2,** and more details on associated traits for each SNP are given in **Supplementary Data**. Most of these SNPs were associated with a large number of traits from a broad range of categories, including BMI/adiposity-related traits and other closely linked traits such as lipid levels. For comparison, we selected the two SNPs which were genome-wide significant in the exposure GWAS we used and had also been identified as associated with vitamin D in a previous GWAS. For each of these SNPs, we conducted the same PheWAS analysis. These SNPs also showed associations with a number of other traits in OpenGWAS, however, examining those traits further showed that the majority were other GWAS studies of vitamin D levels (see **Supplementary Data**). Excluding other vitamin D GWAS left very few associations for those SNPs, all of which were either plausibly on the same causal pathway as vitamin D (e.g. skin colour, childhood sunburn occasions) or were unnamed proteins.

**Table 2:** PheWAS results for SNPs showing evidence of an effect of Vitamin D on BMI.

| SNP | Beta | SE | P-value | No. Traits |
| --- | --- | --- | --- | --- |
| *SNPS SHOWING EVIDENCE OF A CAUSAL EFFECT* |  |  |  |  |
| rs35733741 | -1.719 | 0.154 | 5.94E-29 | 134 |
| rs804280 | 1.067 | 0.130 | 2.92E-16 | 168 |
| rs1972994 | 0.828 | 0.103 | 7.91E-16 | 115 |
| rs1047891 | -0.995 | 0.127 | 4.75E-15 | 156 |
| rs34293138 | 1.155 | 0.153 | 5.22E-14 | 180 |
| rs7148857 | -1.002 | 0.136 | 1.48E-13 | 54 |
| rs2245133 | -0.776 | 0.110 | 1.87E-12 | 21 |
| rs7178572 | -0.642 | 0.124 | 2.38E-07 | 45 |
| rs7930750 | 0.145 | 0.028 | 3.09E-07 | 18 |
| rs7828742 | -0.365 | 0.082 | 8.81E-06 | 35 |
| rs55886116 | -0.580 | 0.145 | 6.33E-05 | 7 |
| rs6773343 | 0.584 | 0.150 | 9.83E-05 | 19 |
| rs2934744 | -0.312 | 0.080 | 1.01E-04 | 222 |
| rs10771090 | 0.590 | 0.159 | 2.11E-04 | 19 |
| rs1011468 | -0.449 | 0.123 | 2.65E-04 | 13 |
| *SNPS IN PREVIOUSLY IDENTIFIED GENE REGIONS* |  |  |  |  |
| rs10859995 | 0.005 | 0.043 | 0.906 | 16 |
| rs8018720 | 0.003 | 0.072 | 0.965 | 11 |

For each SNP, Beta is the Wald ratio effect estimate obtained using only that SNP, Std. Error and P-value are the corresponding standard error and p-values. No. Traits gives the number of other traits (defined as individual GWAS studies) that the SNP was associated with in OpenGWAS. Association was defined as having a p-value of $< 5 \times 10^{-8}$ in a GWAS study in OpenGWAS (excluding that used for vitamin D in this paper). Similar traits were not excluded from this count. A full list of all traits is given in the **Supplementary Material**.

**Figure 2:**
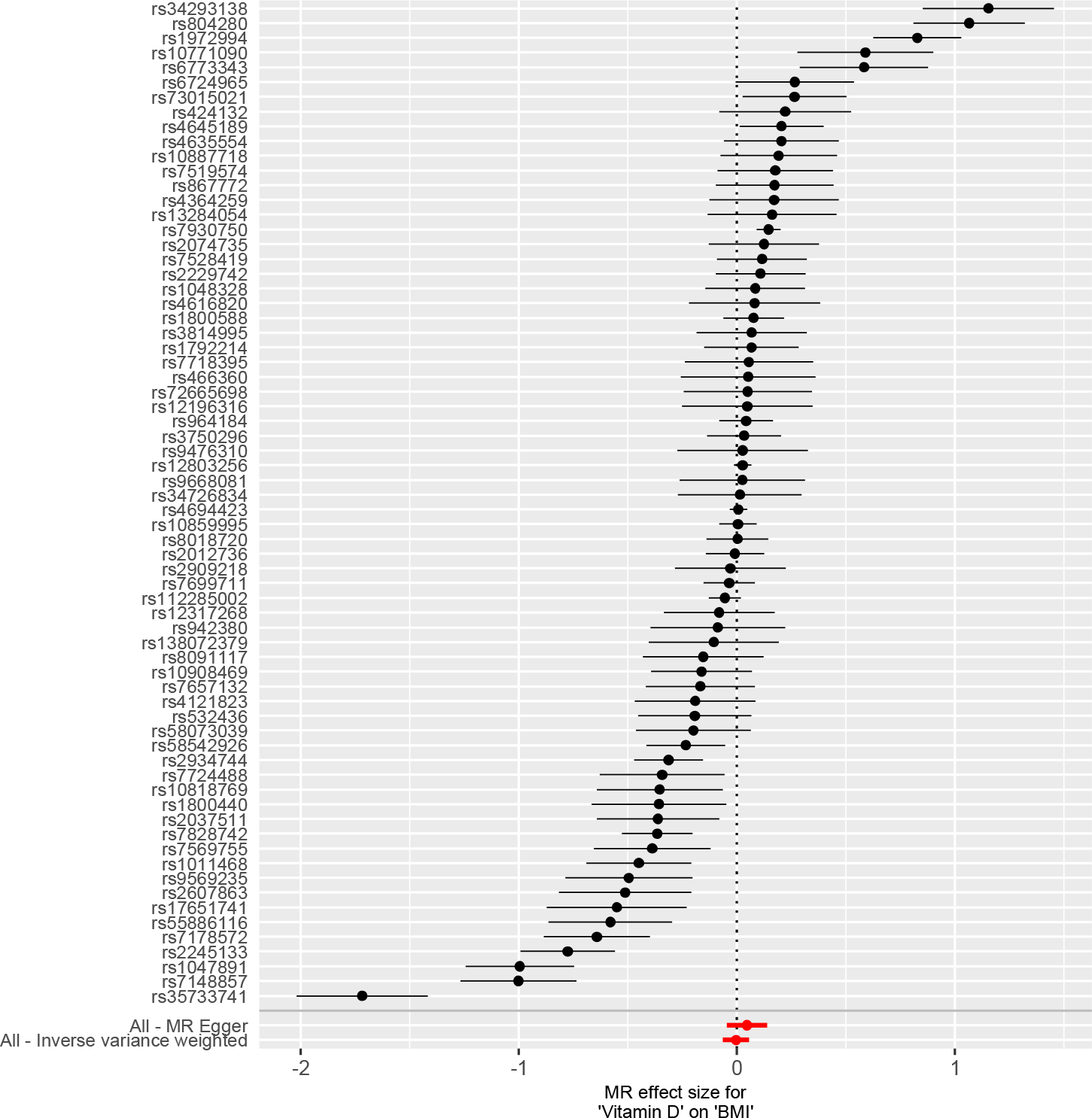
Forest plot of the results from the single SNP Wald estimation Plot shows the estimated effect of Vitamin D on BMI obtained using a Wald ratio for each SNP and the overall estimate obtained from IVW and MR Egger estimation. Number of SNPS; 68

Due to the prevalence of adiposity-related traits in our PheWAS, we further conducted Steiger filtering to remove SNPs that were more strongly associated with BMI than vitamin D.^31^ We also conducted a bi-directional MR of BMI on vitamin D to identify whether the associations of the SNPs identified in the vitamin D GWAS with adiposity related traits were due to a causal effect of adiposity on vitamin D. The second panel in **Table 1** gives the MR effect estimates with Steiger filtering applied to remove SNPs that explained more variation in BMI than vitamin D. Although many of the SNPs which showed associations with BMI in our PheWAS analysis were likely to be pleiotropic, only 2 SNPs were removed by the Steiger filtering and the overall results were not substantively changed.

**Table 3** gives the results for the estimated causal effect of BMI on vitamin D. For this analysis we selected independent SNPs that were genome-wide significant in the GWAS we had previously used as our outcome (p<5×10^−8^, r^2^ < 0.001) as our SNP-exposure association and then extracted the SNP-vitamin D association from the GWAS we had previously used as our exposure. We then applied the same set of MR effect estimates as previously. The results from this analysis show evidence of a causal effect of BMI on vitamin D (IVW: effect of na SD increase in BMI on an SD change in vitamin D: -0.09 SD, 95% CI: [-0.11, -0.07]). This result was consistent across the different pleiotropy robust MR methods applied.

**Table 3:** MR estimates for the effect of BMI on Vitamin D.

|  | **Beta** | **SE** | **P-value** | **95% C.I.** |
| --- | --- | --- | --- | --- |
| IVW | -0.091 | 0.010 | $2.56 \times 10^{-21}$ | [-0.11 -0.07] |
| MR-Egger | -0.092 | 0.025 | $3.29 \times 10^{-4}$ | [-0.14 -0.04] |
| Weighted median | -0.083 | 0.011 | $1.01 \times 10^{-13}$ | [-0.10 -0.06] |
| Weighted mode | -0.093 | 0.023 | $5.88 \times 10^{-5}$ | [-0.14 -0.05] |

MR estimates for the effect of BMI on vitamin D. Number of SNPs: 477. $\beta$ : Estimated SD change in circulating vitamin D per SD increase in BMI, SE: standard error, 95 % C.I.: 95% confidence interval.

## Discussion

Lower vitamin D has been associated with higher BMI and obesity-related diseases in numerous observational studies.7-9 However, there is high potential for unobserved confounding or reverse causation in these studies, and therefore these associations may not be due to a causal effect of vitamin D on BMI. MR is a method for estimating causal effects that is not biased by unobserved confounding that can bias conventional observational studies. A previous MR study showed no effect of vitamin D on BMI, however, that study used a small number of SNPs (4) as genetic instruments and had a much smaller sample size than is now available.^17^ Here we have used two-sample summary data MR, and the latest GWAS study of vitamin D, which identified 69 independent SNPs associated with vitamin D, to more robustly estimate the effect of vitamin D on BMI. Our results show no evidence of a causal effect of vitamin D levels on BMI. Follow up analyses of the individual SNPs show that this absence of an effect is not due to a lack of power in the MR analysis or in the GWAS studies used to identify SNPS. The SNPs that did show evidence of a causal effect were highly pleiotropic and associated with numerous other traits in addition to vitamin D.

Conversely, SNPs for which there was previous biological evidence of the relevance of that SNP in circulating vitamin D levels showed no evidence of a causal effect of vitamin D on BMI and were primarily associated with other vitamin D traits in follow-up PheWAS analysis.

We also conducted a bidirectional MR to estimate the effect of BMI on vitamin D, using the same datasets but with BMI as the exposure and SNPs significantly associated with BMI, at genome-wide significance, used as instruments. This analysis showed evidence of a causal effect of BMI on vitamin D levels that was consistent across a range of pleiotropy robust methods; IVW effect estimate of a SD change in BMI on a SD change in vitamin D: -0.09, 95% CI: [-0.11, -0.07]). This suggests that conventional observational associations of vitamin D with BMI are also likely driven by reverse causation, in addition to unobserved confounding.

We applied Steiger filtering to our estimation of the effect of vitamin D on BMI to remove SNPs that explained more variation in BMI than in vitamin D. However, despite the PheWAS showing that many SNPs were likely to be pleiotropic, and the true direction of the causal effect being from BMI to vitamin D, only 2 SNPs were removed by the filtering. This approach therefore did not substantively change the results obtained or reduce the large number of pleiotropic SNPs from the analysis. Overall, these results show that using the largest GWAS available for a trait, and relying only on statistical approaches for removing pleiotropic or reverse causal SNPs, may not always increase the reliability of the results from the MR study. Where possible, incorporating biological knowledge and external SNP validation can help improve the reliability of MR results. In our results, the direction of the bias for each individual SNP was balanced across the SNPs that were likely to be pleiotropic and so did not appear to bias the overall result, however, this will not hold in all cases.

## Limitations

There are, however, several limitations to this study. The data underlying this analysis were all based on a UK population with white European ancestry. It is important therefore to explore whether the result would hold in other regions and ancestry groups. Additionally, if the effect of vitamin D levels on BMI are non-linear across different levels of vitamin D, then the results obtained in this study would not be generalisable to other populations with different vitamin D levels. The non-linearity in the effects of vitamin D has been suggested in other MR studies^33-35^, however this is an area of ongoing debate and methodological challenges.^36-38^ As we had summary-level data for this study, we were unable to explore any non-linearity in effects.

The largest GWAS available for both our exposure and outcome included UK Biobank as the majority/all of their sample. This increases the risk of ‘winners curse’ bias in our results. However, we had strong instruments for our analysis which limits the potential for incorrect inference from this bias.^39^ We were also unable to explore violations of the second IV assumption. Our results showed that a number of the SNPs included in the analysis violated the third IV assumption, but the effects of these SNPs on the overall result were balanced. However, violations of the second IV assumption can occur due to population stratification, dynastic effects or assortative mating which may affect all SNPs and so not have been detected by our approach. ^26^ This bias can be avoided by using within family MR, however, this requires large GWAS samples of siblings or trios which are often not available. ^40^

## Conclusions

In this study, we have used the latest and largest GWAS of vitamin D, and multiple methodological approaches, to more robustly estimate the causal effect of vitamin D on BMI. Our results support an overall conclusion that the commonly observed association between higher vitamin D and lower BMI in conventional observational studies is due to a reverse causal effect of BMI on vitamin D, and potentially other confounding factors, but is unlikely due to a causal effect of vitamin D on BMI. Targeting vitamin D is therefore unlikely to be an effective strategy for reducing adiposity.

## Data Availability

The data used in this study is available at https://gwas.mrcieu.ac.uk/.

## Contributions

MC conducted the analysis and wrote the first draft of the paper. JB and ES supervised the analysis and contributed to the writing and editing of the paper.

## Data availability

All analysis was conducted using publicly available data accessed through the IEU OpenGWAS infrastructure.

## Code availability

All analyses were conducted in R version 4.2.2, The code used to generate these results has been archived at (https://github.com/eleanorsanderson/VitD_BMI).

