## Supplementary figure for "Estimating the effect of circulating vitamin D on body mass index: a Mendelian randomization study"

**SUPPLEMENTARY MATERIAL**

Minal Chadha^1^, Joshua Bell^1,2^ and Eleanor Sanderson^1,2^

1. Population Health Sciences, Bristol Medical School, Bristol University, Bristol, UK.
2. MRC Integrative Epidemiology Unit, Bristol University, Bristol, UK.

***Supplementary Figure 1: MR estimates for the effect of vitamin D on BMI***

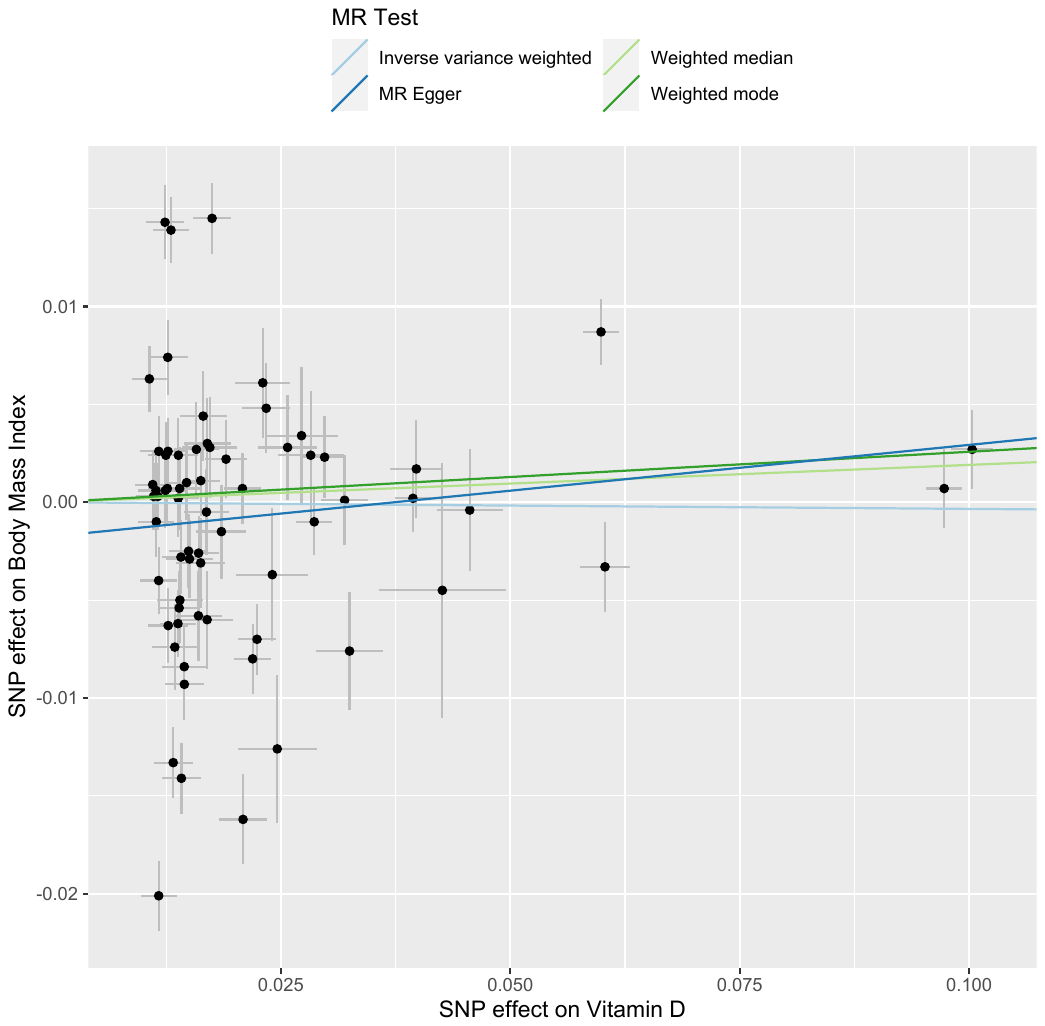

Plot of SNP – vitamin D association (x – axis) against SNP – BMI association (y – axis) for all SNPs used in the MR estimation of the effect of vitamin D on BMI. Plotted lines give the effect estimates using each of the different MR estimators.

***Supplementary Table 1: Single SNP Wald ratios***

| **SNP** | **beta** | **Std. err.** | **p-value** |
| --- | --- | --- | --- |
| rs34293138 | 1.155 | 0.153 | 5.22E-14 |
| rs804280 | 1.067 | 0.130 | 2.92E-16 |
| rs1972994 | 0.834 | 0.103 | 5.02E-16 |
| rs10771090 | 0.590 | 0.159 | 2.11E-04 |
| rs6773343 | 0.584 | 0.150 | 9.83E-05 |
| rs6724965 | 0.266 | 0.139 | 5.57E-02 |
| rs73015021 | 0.265 | 0.122 | 2.94E-02 |
| rs424132 | 0.222 | 0.154 | 1.49E-01 |
| rs4645189 | 0.205 | 0.098 | 3.69E-02 |
| rs4635554 | 0.205 | 0.134 | 1.26E-01 |
| rs10887718 | 0.192 | 0.136 | 1.58E-01 |
| rs7519574 | 0.177 | 0.135 | 1.92E-01 |
| rs867772 | 0.173 | 0.137 | 2.07E-01 |
| rs4364259 | 0.171 | 0.152 | 2.61E-01 |
| rs13284054 | 0.162 | 0.151 | 2.82E-01 |
| rs7930750 | 0.145 | 0.028 | 3.09E-07 |
| rs4616820 | 0.145 | 0.154 | 3.47E-01 |
| rs2074735 | 0.125 | 0.128 | 3.31E-01 |
| rs7528419 | 0.116 | 0.105 | 2.71E-01 |
| rs2229742 | 0.109 | 0.105 | 3.00E-01 |
| rs1048328 | 0.085 | 0.117 | 4.67E-01 |
| rs1800588 | 0.077 | 0.071 | 2.73E-01 |
| rs3814995 | 0.068 | 0.129 | 5.99E-01 |
| rs1792214 | 0.068 | 0.111 | 5.41E-01 |
| rs7718395 | 0.055 | 0.150 | 7.13E-01 |
| rs466360 | 0.053 | 0.158 | 7.39E-01 |
| rs72665698 | 0.050 | 0.150 | 7.39E-01 |
| rs12196316 | 0.048 | 0.153 | 7.52E-01 |
| rs964184 | 0.043 | 0.063 | 4.97E-01 |
| rs3750296 | 0.034 | 0.086 | 6.97E-01 |
| rs9476310 | 0.027 | 0.153 | 8.60E-01 |
| rs12803256 | 0.027 | 0.020 | 1.77E-01 |
| rs34726834 | 0.014 | 0.145 | 9.20E-01 |
| rs9668081 | 0.009 | 0.147 | 9.53E-01 |
| rs4694423 | 0.007 | 0.021 | 7.26E-01 |
| rs10859995 | 0.005 | 0.043 | 9.06E-01 |
| rs8018720 | 0.003 | 0.072 | 9.65E-01 |
| rs2012736 | -0.007 | 0.068 | 9.23E-01 |
| rs7699711 | -0.024 | 0.059 | 6.81E-01 |
| rs2909218 | -0.030 | 0.130 | 8.20E-01 |
| rs112285002 | -0.055 | 0.038 | 1.51E-01 |
| rs12317268 | -0.081 | 0.130 | 5.32E-01 |
| rs942380 | -0.087 | 0.157 | 5.79E-01 |
| rs138072379 | -0.106 | 0.153 | 4.89E-01 |
| rs8091117 | -0.154 | 0.141 | 2.76E-01 |
| rs10908469 | -0.162 | 0.118 | 1.71E-01 |
| rs7657132 | -0.167 | 0.127 | 1.88E-01 |
| rs4121823 | -0.191 | 0.141 | 1.78E-01 |
| rs532436 | -0.193 | 0.133 | 1.47E-01 |
| rs58073039 | -0.198 | 0.135 | 1.41E-01 |
| rs58542926 | -0.234 | 0.092 | 1.13E-02 |
| rs2934744 | -0.312 | 0.080 | 1.01E-04 |
| rs7724488 | -0.342 | 0.145 | 1.86E-02 |
| rs10818769 | -0.354 | 0.147 | 1.64E-02 |
| rs1800440 | -0.357 | 0.157 | 2.30E-02 |
| rs2037511 | -0.362 | 0.144 | 1.17E-02 |
| rs7828742 | -0.365 | 0.082 | 8.81E-06 |
| rs7569755 | -0.388 | 0.136 | 4.48E-03 |
| rs1011468 | -0.449 | 0.123 | 2.65E-04 |
| rs9569235 | -0.495 | 0.149 | 9.14E-04 |
| rs2607863 | -0.512 | 0.154 | 9.14E-04 |
| rs17651741 | -0.550 | 0.164 | 7.69E-04 |
| rs55886116 | -0.580 | 0.145 | 6.33E-05 |
| rs7178572 | -0.642 | 0.124 | 2.38E-07 |
| rs2245133 | -0.776 | 0.110 | 1.87E-12 |
| rs1047891 | -0.995 | 0.127 | 4.75E-15 |
| rs7148857 | -1.002 | 0.136 | 1.48E-13 |
| rs35733741 | -1.719 | 0.154 | 5.94E-29 |

Individual SNP Wald ratios for the effect of vitamin D on BMI for each SNP selected as an instrument for vitamin D. b is the Wald ratio effect estimate obtained using only that SNP, Std. Err and p-value are the corresponding standard error and p-value.
